# In-phase bilateral upper limb exercises improve cognitive and motor function in Progressive Multiple Sclerosis: A pilot randomized controlled trial

**DOI:** 10.64898/2025.11.24.25340607

**Authors:** Dimitris Sokratous, Charalambos Costa Charalambous, Marios Pantzaris, Kyriaki Michailidou, Nikos Konstantinou

## Abstract

**Introduction:** Progressive Multiple Sclerosis impairs both motor and cognitive functions and reduces quality of life. While complex goal-directed movements are challenging due to cognitive deficits, in-phase bilateral exercise require less attentional demand and cognitive effort. Thus, this type of exercise may improve motor function and cognition in this cohort. Previous studies in people with Progressive Multiple Sclerosis indicated a strong connection between cognitive function and upper limb function, yet whether in-phase bilateral exercises can improve cognition and motor performance is still unclear. In this trial, we aimed to examine the effect of in-phase bilateral upper limb exercises on the cognition, motor function and quality of life in people with Progressive Multiple Sclerosis.

**Methods:** Twenty people (11 females and 9 males, mean age = 55.8) with Progressive Multiple Sclerosis were randomly allocated (1:1) to either an experimental or active control group in a 12-weeks intervention. The experimental group performed in-phase bilateral upper limb exercises (total of 36 sessions), while the active control group followed a conventional exercise program (total of 12 sessions). A repeated measures ANOVA analysis was conducted to determine significant differences in information processing speed, manual dexterity, gait, balance, fatigue and quality of life.

**Results:** Post-hoc analyses revealed that the experimental group demonstrated significantly greater improvements than the active control group in information processing speed (t(18) = 8.6, *p* < 0.05) and across all secondary outcome measures (all *p*s < 0.05).

**Conclusion:** These findings indicate that in-phase bilateral exercise program, which require less cognitive effort than other types of bilateral goal-directed coordination, can enhance information processing speed, motor functions, fatigue and quality of life in people with Progressive Multiple Sclerosis.

## Introduction

Multiple sclerosis (MS) is the most common inflammatory and neurodegenerative disease of the central nervous system (1). Given that most people with Relapsing-Remitting Multiple Sclerosis eventually transit to the Progressive MS (PMS), which includes both both Secondary Progressive MS and Primary Progressive MS (1), understanding the progression and treatment options for PMS becomes critical. This transition is associated with an increase in disability and a steady accumulation of neurological impairment.

People with PMS (pwPMS) not only experience physical impairment but also commonly suffer from cognitive dysfunction (2), which significantly affects their quality of life (QoL). Among cognitive domains, information processing speed is one of the most commonly impaired (3,4) in both Secondary and Primary Progressive MS (5). Various cognitive rehabilitation programs (4,6) have shown efficacy in improving MS-related cognitive dysfunctions, such as learning, memory, attention and cognitive processing.

Exercise has been shown to improve both cognitive and motor functions (7,8), as well as overall QoL in people with MS (9), offering a holistic intervention compared to cognitive training alone (10). Evidence from healthy individuals and people with MS indicates a close relationship between cognitive function and upper limb performance (11,12), supported by dense neural projections linking the anterior cingulate cortex, motor cortex and spinal pathways (13). Impaired information processing speed contributes to reduced manual dexterity, limiting independence and daily functioning (12,14).

Emerging evidence indicates a close interdependence between motor control and cognitive function (13,15,16). Within this neurofunctional framework, bilateral upper limb movements, particularly in-phase bilateral movements, where both limbs move simultaneously in the same direction, have emerged as a promising intervention for enhancing cognitive performance. These movements engage both hemispheres, promoting interhemispheric communication via the corpus callosum and strengthening connectivity between the supplementary motor area and the primary motor cortex (13,17,18). These neural adaptations facilitate more efficient integration of sensory and motor information, leading to improvements in information processing speed, a key determinant of higher-order cognitive functions such as attention, working memory and executive control (19–23).

Importantly, in-phase bilateral movements require lower attentional and motor control demands than anti-phase or unilateral patterns, making them particularly suitable for rehabilitation in populations with cognitive or motor impairments, such as pwPMS (24–26). Preliminary evidence from a clinical trial in people with Relapsing-Remitting MS (27) has shown that in-phase bilateral upper limb exercises significantly improve information processing speed, further supporting their potential as a targeted intervention to enhance both cognitive and motor outcomes in PMS.

The cognitive benefits of exercise may be further enhanced through group-based circuit training, which integrates physical, cognitive and social components. Group exercise not only provides structured physical conditioning but also fosters social interaction, enhancing motivation and engagement while activating brain regions involved in social cognition and executive control (28,29). The interactive nature of group training creates an engaging environment that promotes adherence and exercise intensity, both key determinants of cognitive improvement (30). Moreover, structured group exercise programs, particularly those implemented in circuit formats, have been associated with greater neuroplasticity, improved brain function (29) and reduced risk of cognitive decline (28). Circuit training, characterized by rapid transitions between different exercises, delivers simultaneous physical and cognitive stimulation. Within this framework, the motor-cognitive model proposed by Herold et al. (2018) suggests that embedding cognitive demands, such as task switching, attentional control and working memory, into movement optimally engages prefrontal and cerebellar networks, thereby enhancing executive functions (31).

Despite evidence for dual-task motor and cognitive training in pwPMS (7,32), it remains uncertain whether specific low-cognitive-demand exercise modalities can improve cognitive and motor functions comparably. This is particularly relevant for patients with cognitive impairments who may struggle to follow complex dual-task instructions. The present pilot study therefore implemented a structured, group-based circuit training intervention incorporating in-phase bilateral upper limb exercises, designed to enhance information processing speed, manual dexterity, gait, balance, fatigue and QoL in pwPMS.

## Materials and Methods

### Participants

Participants were recruited from The Cyprus Institute of Neurology and Genetics in September 2023. All participants underwent a neurological evaluation to confirm eligibility prior to enrollment. A total of 28 pwPMS consented to participate, however, eight did not meet the eligibility criteria and were excluded (Fig. 1), yielding a final sample of 20 participants. Participant allocation was completed between September 25 and 30, 2023, following a review of medical records. Only the neurologist had access to identifying data and was not informed of the participants’ group assignments. The study was conducted between October 2 and December 22, 2023, and was registered at ClinicalTrials.gov (NCT06436131) with ethical approval obtained from the Cyprus National Bioethics Committee (EEBK/EΠ/2022/32).

**Fig 1.** CONSORT Flow Diagram of Study Participants. Eight participants did not meet the inclusion criteria, therefore were not enrolled in the study. Enrolled participants were randomized to experimental (n=10) and active control (n=10) group. All of the participants completed the study and none of them was excluded from the data analysis.

The inclusion criteria included 1) diagnosis of PMS (primary or/and secondary PMS), 2) Expanded Disability Status Scale (EDSS) (33) score between three and six, 3) no relapse within the last 30 days, 4) aged between 30 and 70 years and 5) Mini Mental State Examination (34) score between 20 and 30 (mild to no cognitive impairment). The exclusion criteria included 1) history of any disease affecting the central nervous system other than MS (e.g., stroke), 2) history of cardiovascular disease (e.g., myocardial infarction), 3) severe orthopedic disorders (e.g., knee/hip replacement), 4) mental disorders (e.g., depression), 5) pregnancy during the implementation of the study timeline, 6) hearing impairments (i.e., deafness), 7) visual deficit (e.g., optic neuritis), 8) history of epileptic seizures and 9) spasticity level on upper or lower limbs more than 1+ (slight increase in muscle tone) according to Modified Ashworth Scale (35).

Additionally, participants were advised to continue their usual prescribed medication throughout the study duration and they were advised to continue their usual routine and avoid receiving any other exercise program during the study. Furthermore, all participants read and signed a written informed consent, while all procedures were approved and conducted in accordance with the ethical guidelines of the Cyprus National Bioethics Committee before recruitment.

#### Study design

This double-blind, two-arm randomized controlled trial aimed to evaluate the effects of a 12-week in-phase bilateral upper limb exercise program on information processing speed in pwPMS. Participants were randomly assigned to either an experimental or an active control group (Fig 2).

**Fig 2.** Timeline and Schematic Representation of the Study Design. B, Baseline; I, intervention; c, clinical assessments; q, questionnaires; *, assessment points for each group. The first two rows represent the experimental group (white row) and the active control group (black row). The third and fourth rows (grey) indicate the timing of assessments and corresponding study weeks. The study spanned a total of 15 consecutive weeks, comprising a 3-week baseline phase followed by a 12-week intervention phase. During the intervention phase, participants in the experimental group engaged in an exercise protocol emphasizing in-phase bilateral upper limb movements. In contrast, participants in the active control group completed a conventional exercise regimen.

The experimental group received supervised training from a certified fitness instructor, whereas the active control group was supervised by a physiotherapist, both blinded to group allocation. Clinical assessments for both groups were conducted by an independent physiotherapist blinded to group allocation. To minimize intergroup contamination, training sessions for the two study groups were conducted separately. Randomization was conducted using computer-generated randomization, stratified by EDSS score, age, gender and hand dominance (Table 1), factors known to influence exercise performance (36–38) and relevant to the outcome measures.

**Table 1.**
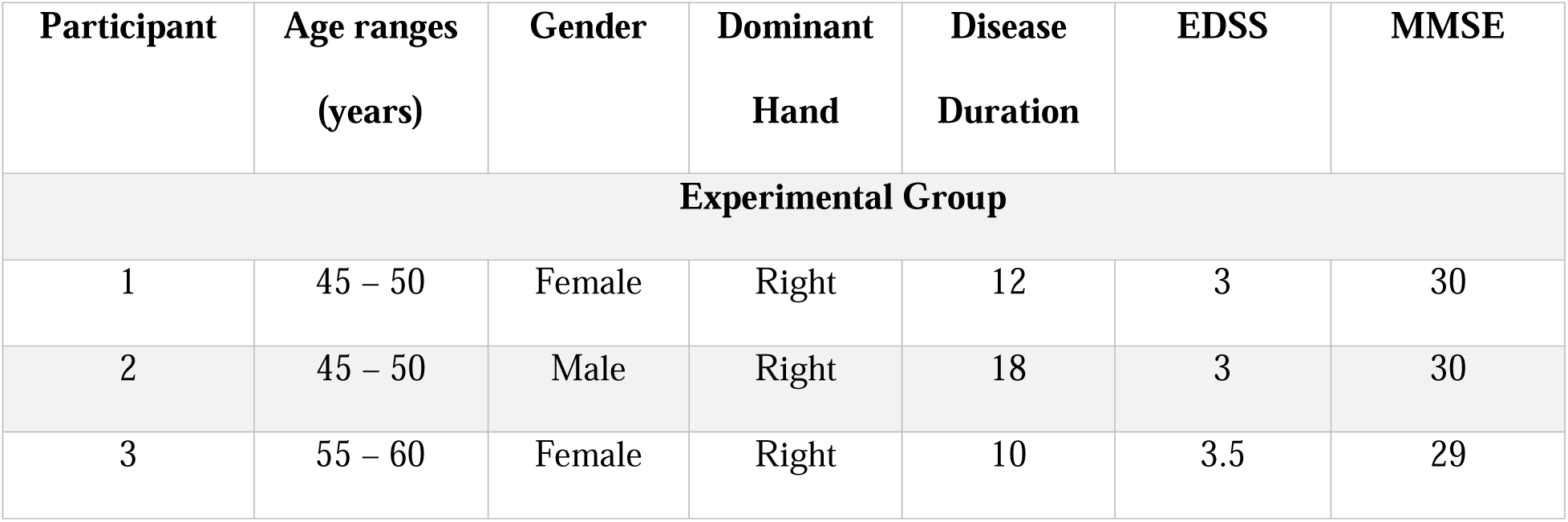
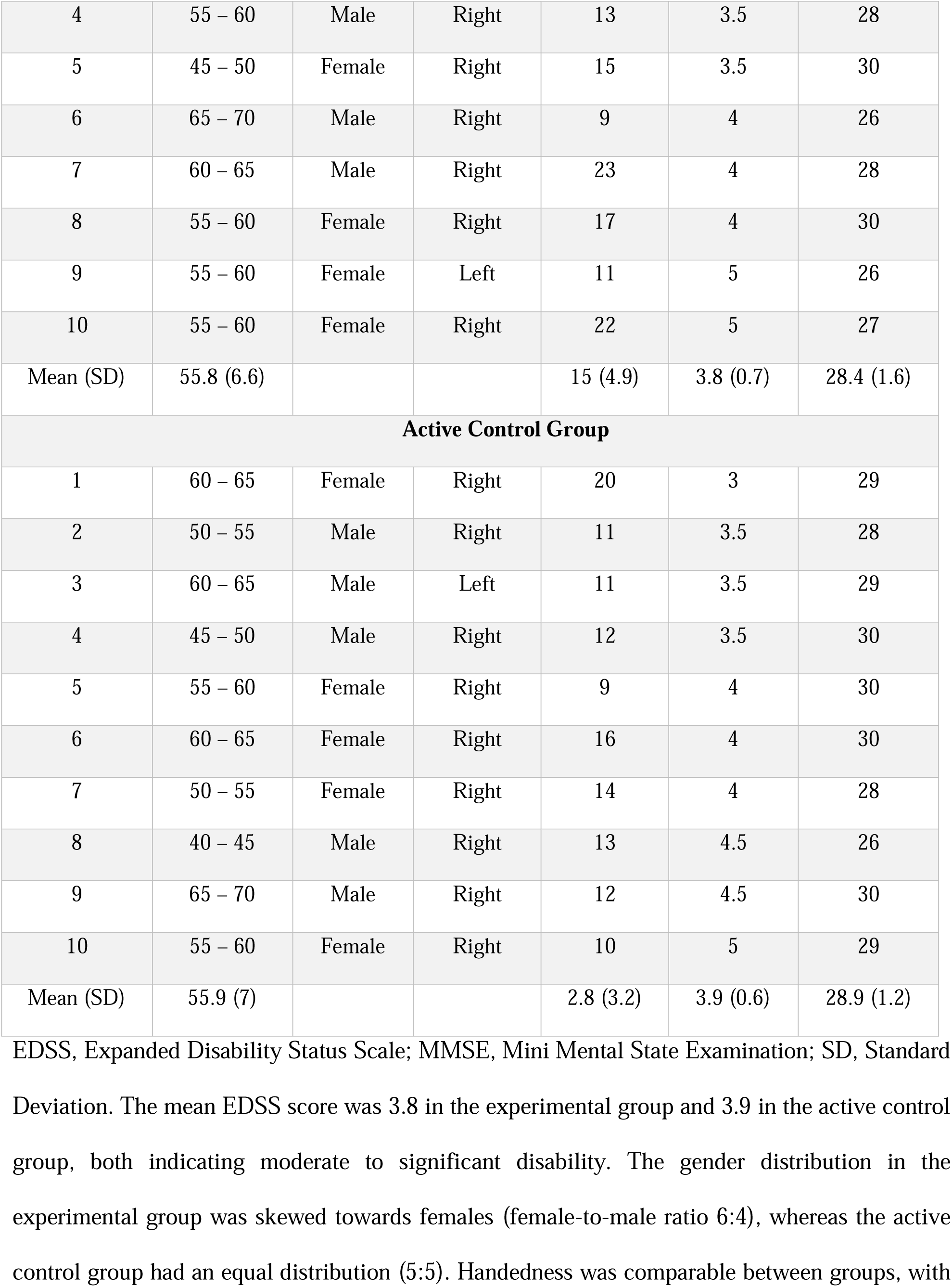
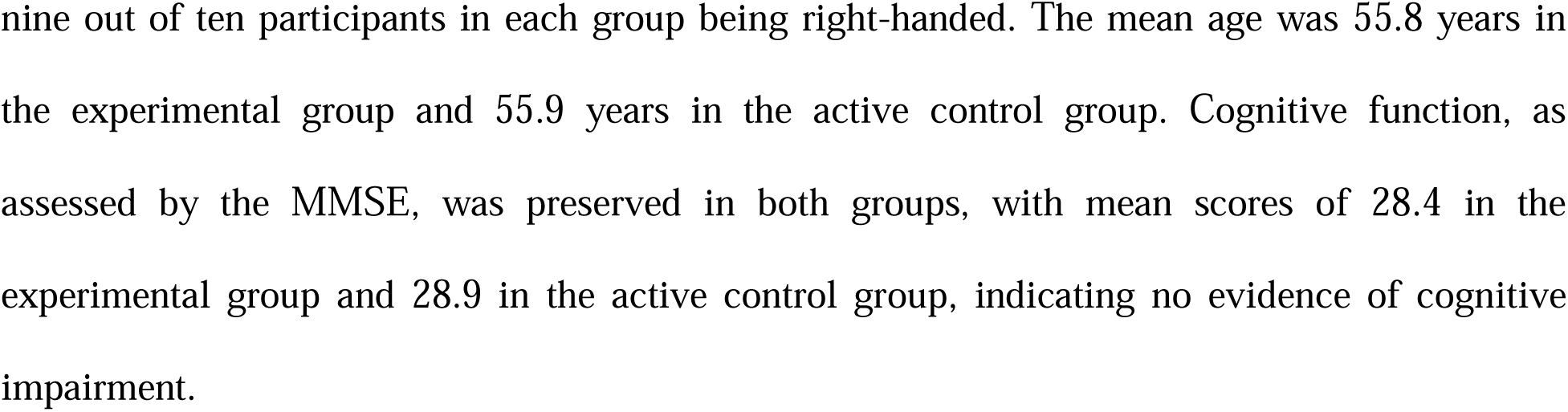
Participants’ Demographic and Baseline Clinical Characteristics.

#### Baseline phase

All participants entered a three-week baseline phase that began simultaneously for the entire cohort, with assessments conducted once per week. Cognitive and motor outcomes were recorded at each time point and two subjective questionnaires (see outcome measures) were administered during the final week of the baseline phase (Fig 2).

#### Intervention phase

The 12-week intervention included three sessions per week for the experimental group and one session per week for the active control group. The control group exercised once weekly to reflect the level of physical activity typically recommended by physiotherapists within the national health system for the chronic management of these conditions and for maintaining general health and QoL. For ethical reasons, a completely inactive control group was not included. This design allowed us to examine whether the specific exercise protocol conferred additional benefits beyond those expected from standard guideline-based activity levels.

##### Experimental Grοup

Every intervention session consisted of a five minute warm-up (i.e., whole body range of motion exercises), followed by 40min to 50min of the main in-phase bilateral upper limb exercise protocol described below and a cool down for five minutes (i.e., passive stretching exercises of the muscle groups which are involved in the main part). Throughout the intervention phase, each participant underwent three clinical assessments and completed two subjective questionnaires, resulting in three distinct data points per participant over the course of the intervention (Fig. 2). To be included in the analysis, participants were required to attend at least 27 out of 36 (i.e., 75%) of the total sessions for their study group (27,39).

Exercises were conducted in a sports hall, involving in-phase bilateral upper limb movements in a group circuit training, adapted from MS guidelines (40) and previous studies in Relapsing-Remitting MS (27). Each session included three sets of nine exercises targeting large upper-limb muscle groups (e.g., shoulder: flexors, extensors, rotators, abductors, adductors, horizontal abductors and adductors; elbow: flexors and extensors). Three additional exercises targeted large lower-limb muscle groups (e.g., hip: flexors, extensors, abductors, adductors; knee and ankle flexors and extensors). Lower-limb exercises were performed between upper-limb exercises to allow muscle relaxation. Each exercise lasted one minute, during which participants performed as many repetitions as possible. A two-minute rest period was provided between sets.

The program incorporated sports-based technical skills, such as basketball (e.g., passing, catching and throwing) and volleyball (e.g., passing and receiving). Fitness exercises included diagonal movements of proprioceptive neuromuscular facilitation technique (41) using resistance bands and open-chain upper-limb exercises performed with 1kg dumbbells (e.g., shoulder flexion/extension or shoulder abduction/adduction with extended elbows) (Table 2). To maintain participants’ engagement and ensure that the individual Rate of Perceived Exertion remained within the target range of three to six, the exercise protocol was progressively adapted over the 12-week intervention period through systematic adjustments to the level of difficulty. Specifically, elastic bands with varying resistance levels, dumbbells of different weights and modifications in passing distances during ball exercises were introduced. All sessions were conducted in a temperature-controlled environment (24 °C) under standardized safety conditions (e.g., mats) to minimize the risk of falls. This approach ensured that the exercises remained challenging yet achievable, maximizing the benefits for participants without risking overexertion.

**Table 2.**
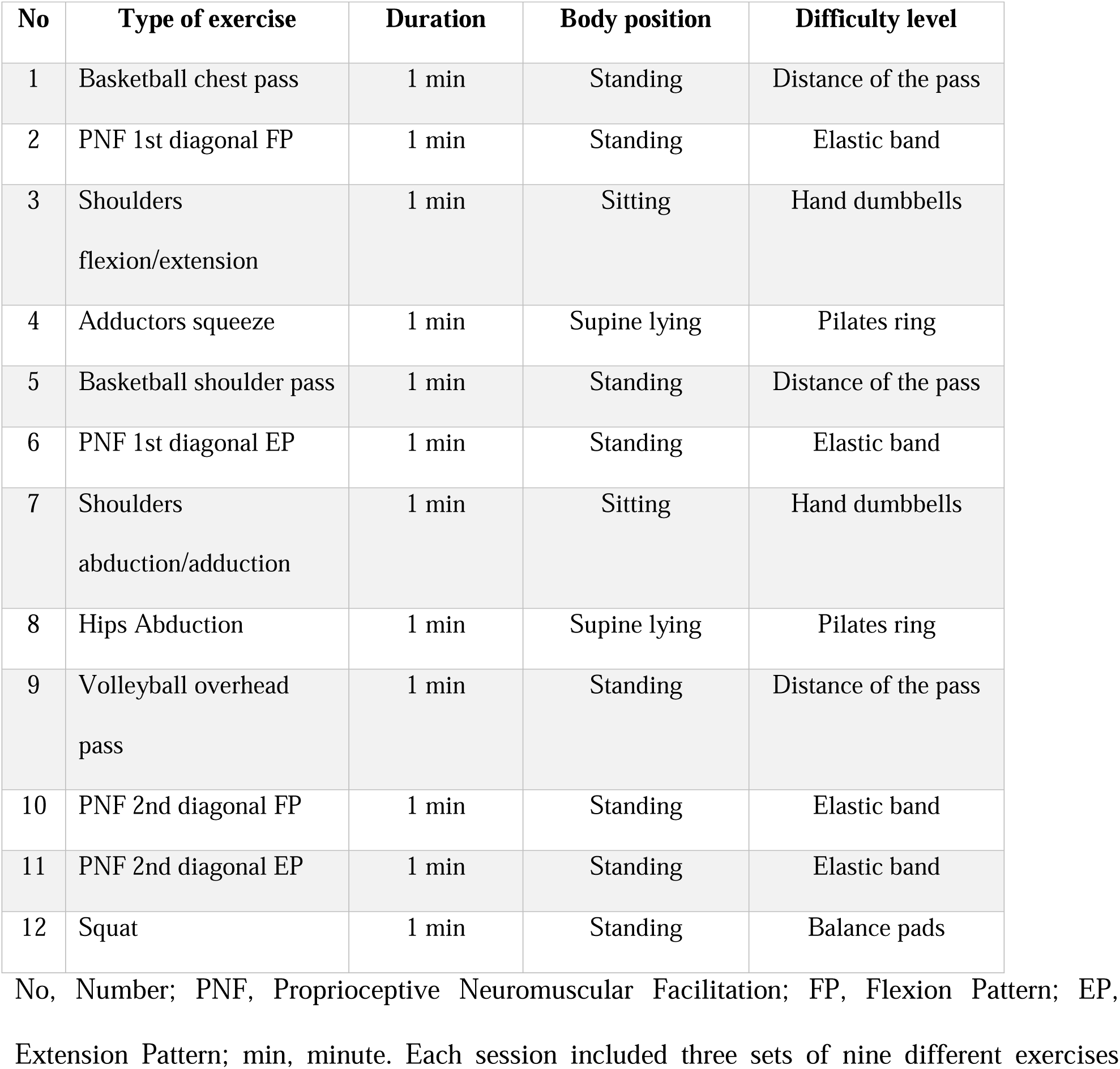
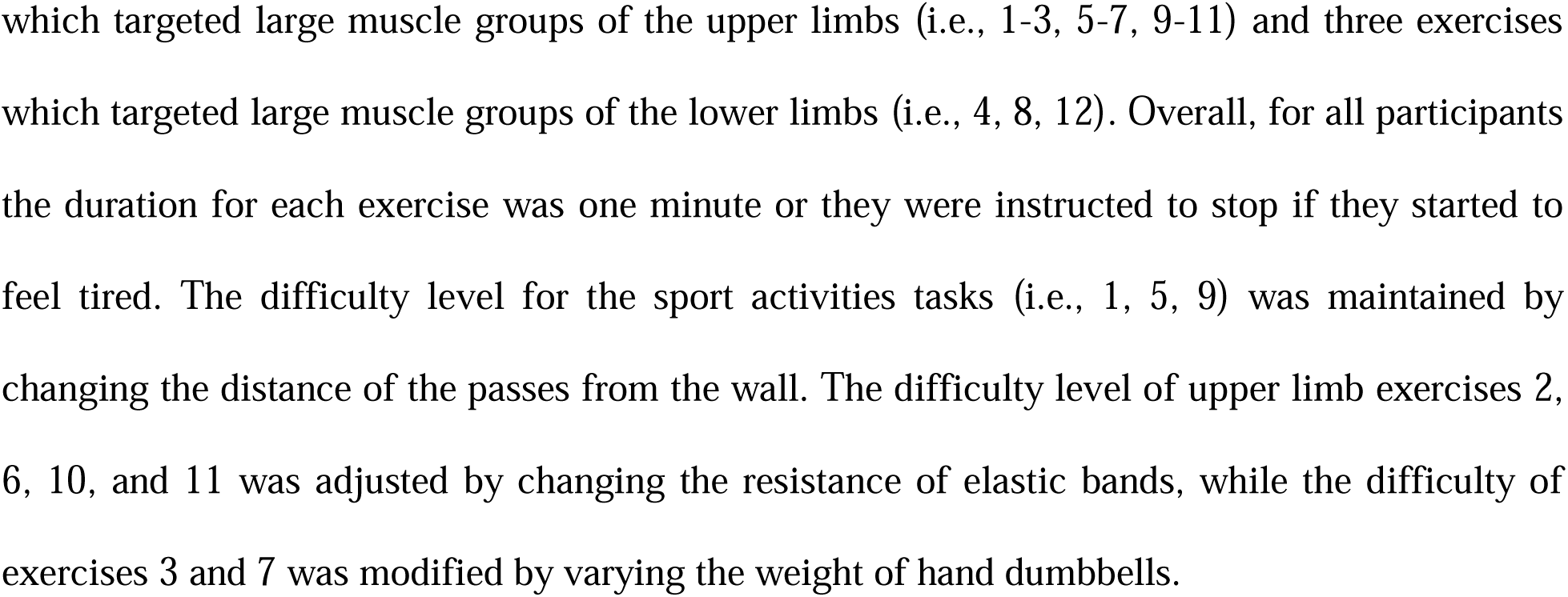
Overview of the Exercise Protocol.

##### Active Control Group

All exercise sessions and clinical assessments were conducted at the Physiotherapy Unit of the Cyprus Institute of Neurology and Genetics. Participants in the active control group attended one-on-one sessions with the same physiotherapist. Participants were required to complete at least 9 out of 12 sessions (i.e., 75%) for their data to be included in the analysis (27,39). Every session consisted of a five minute warm-up (i.e., whole body range of motion exercises), followed by 40min to 50min of the main conventional exercise program described below and a cool down for five minutes (i.e., passive stretching exercises of the muscle groups which are involved in the main part).

The conventional program was implemented according to previously published protocols and consisted of strengthening exercises for the major trunk muscle groups (42), resistance exercises for the upper limbs and treadmill training (43).

###### Trunk Strengthening Exercises

The trunk-strengthening component consisted of exercises targeting the flexor (rectus abdominis) and extensor (erector spinae) muscles. During each exercise, participants held the final static position for three seconds and performed five to ten repetitions, depending on individual tolerance. A three-second pause was allowed between repetitions, with a maximum of one-minute rest between exercises. The difficulty level increased gradually by extending the hold time and increasing the number of repetitions, based on participants’ tolerance.

###### Upper-Limb Strengthening Exercises

The upper-limb strengthening component included exercises targeting the flexor, extensor, internal and external rotator muscle groups of the shoulder joint, as well as the flexor and extensor muscle groups of the elbow joint. Each exercise was performed for five to ten repetitions and one to three sets, depending on individual tolerance. A three-second pause was allowed between repetitions, and a maximum of one-minute rest was provided between sets. The difficulty was gradually increased by either increasing the number of repetitions or the resistance of the elastic band, based on participants’ capabilities.

###### Aerobic Exercises

To complete the session, all participants performed 10 – 15min of treadmill walking at a pace of 1.5 – 2.5 km/h or cycling on a static cycle ergometer (MOTOmed Loop Parkinson) at 20 – 40 bpm. The intensity of these aerobic exercises was adjusted to individual tolerance.

### Outcome Measures

Previous studies have indicated that information processing speed is the most common cognitive deficit in pwPMS (3,4) and it is correlated with manual dexterity (14,44). Therefore, this clinical trial examined whether a specific exercise protocol, based on in-phase bilateral upper limb exercises, led to greater improvement compared to the minimum exercise recommendation of the national health system.

Cognitive processing and manual dexterity are closely intertwined, with each influencing the other. Several tasks requiring manual dexterity often engage cognitive processes, such as attention, memory and problem-solving, suggesting a bidirectional relationship. This connection is particularly relevant to the current study, as our exercise protocol includes in-phase bilateral upper limb exercises. By targeting these motor skills, we anticipate an improvement in information processing speed, as the two are closely connected. Consequently, we have chosen information processing speed as our primary outcome measure, expecting it to be directly impacted by the effects of manual dexterity improvement, achieved through the in-phase bilateral upper limb exercise protocol.

To ensure methodological consistency the same physiotherapist collected all data by performing the same methodological procedures in a quiet room, across all participants and across all time points, for both the experimental and active control group. To prevent any decline in performance due to participant fatigue, all assessments were conducted between 9 a.m. and 11 a.m. in the same sequence as it is described below.

### Primary Outcome Measure

The primary outcome measure was information processing speed, assessed using the Symbol Digit Modalities Test (SDMT), a widely used and validated tool in people with MS (3). The oral version of the SDMT was employed, in which participants were provided with a sheet displaying nine symbols, each paired with a corresponding number at the top of the page, referred to as the “key.” For example, the symbol “O” might be paired with the number “6,” so the correct response would be “six.” The remainder of the page contained a randomized sequence of these symbols. Participants were instructed to verbally identify the number corresponding to each symbol as quickly and accurately as possible over a two-minute period. The total score was calculated by subtracting the number of errors from the number of items completed. To minimize practice effects across multiple assessments, six alternate forms of the SDMT were created, one for each assessment point, with the order of symbols and corresponding key numbers rearranged (45).

### Secondary Outcome Measures

Secondary outcomes included manual dexterity, evaluated with the Purdue Pegboard Test (PPT) (46) changes in gait speed assessed by the Timed 25-Foot Walk Test (47), walking ability, balance and lower limbs coordination during gait evaluated by the Six Spot Step Test (48). Since fatigue and QoL are key factors affecting people with MS, additional secondary outcome measures were included. These were the Modified Fatigue Impact Scale, a subjective questionnaire assessing the effects of fatigue (49) and the Medical Outcomes Study Questionnaire Short Form 36 Health Survey, which is a tool for evaluating Health-Related QoL (50).

#### Medical Outcomes Study Questionnaire Short Form 36 Health Survey

This is a set of generic, coherent and easily administered QoL subjective questionnaire (51). There are 11 questions in the specific questionnaire administered by the assessor, with 36 items in total covering eight domains scaled from 0 to 100. Higher values indicate better health status. The eight domains include: general health, vitality, physical function, role physical, bodily pain, role emotional, social functioning and mental health. Participants need between 5 to 10 minutes to complete the questionnaire.

#### Modified Fatigue Impact Scale

It is a subjective questionnaire describing the effects of fatigue during the past four weeks (49). The Modified Fatigue Impact Scale consists of 21 questions, which are rated from “0” (low rate) to “4” (high rate) and it is divided into three subscales (i.e., physical, cognitive, and psychosocial). The assessor records the total score of the test as the final test result. A higher score indicates greater impact of fatigue in individuals’ daily life.

#### Purdue Pegboard Test

This is a standardized test of manual dexterity (46). It consists of four subtests, performed on a board in which pins, washers and collars are placed by the participants into two parallel columns of holes, according to the subtest task. The first two subtests are unimanual tasks, which measure dexterity of the right and left hand, respectively. The third subtest is a synchronous bimanual task that requires simultaneous use of both hands to grasp and place the pins. In the fourth subtest, participants perform alternating movements of both hands to complete assemblies of different types of pegs. The score is calculated based on the number of pegs inserted in 30 seconds for the first three subtests, and in 1 minute for the fourth subtest.

#### Timed 25-Foot Walk

It is a quantitative assessment for mobility and lower limb function (47). Participants are directed to one end of a marked 25-foot path and are instructed to walk as quickly as possible. The time is recorded from the start and ends when participants reach the 25-foot mark. The same task is immediately run again by having the participants walk back the same distance. As our participants may be using assistive devices for walking, they were instructed to use them for safety reasons. The final score was the mean score from the two completed trials.

#### Six Spot Step Test

It is a measure replicating a complex range of sensorimotor functions, such as lower limb strength, spasticity, coordination, as well as dynamic balance (48). It is a timed walking test that involves kicking over a number of targets placed along a 5-meter path. The specific test is cognitive demanding, that also includes coordination and dynamic balance. The final score was the mean time of the four runs (52).

##### Analysis plan

###### Within-group analysis

For each outcome measure we calculated the mean values from each time point (separately for baseline and intervention). To investigate the effect of the exercise program on each of the outcome measures, separately for each group, we calculated the differences between phases’ mean values. These differences reflect the degree of the intervention-elicited change on the clinical condition of all participants.

###### Between-groups analysis

To detect if there is a significant effect between the two study groups, comparisons were made between the difference of improvement between the two groups, for each outcome measure. The difference of improvement from each group was calculated by the difference between baseline and intervention mean values of each outcome variable.

### Statistical analysis

Normality and sphericity assumptions were evaluated and appropriate adjustments to the tests were made. If violated, non-parametric Wilcoxon signed-rank test (Greenhouse-Geisser) was used, otherwise the change scores (Intervention – Baseline) were compared between groups using repeated measures ANOVA. Significant effects were followed by Bonferroni-corrected post-hoc *t*–tests. Pearson correlations assessed associations between SDMT and PPT scores using intervention data; significance was Bonferroni-adjusted (0.05/number of tests). The null hypothesis posited no between–group differences in outcome changes and was rejected at adjusted *p*□<□0.05. Analyses were conducted in JASP 0.19.1 (https://jasp-stats.org/).

## Results

Following our sampling plan, a total of twenty participants were enrolled and allocated to the experimental (n=10) and the active control (n=10) groups. All participants completed the exercise program without complaints or side effects. As described in detail below, results indicated that participants from the experimental group had on average greater improvement when compared with the active control group on all outcome measures. Data are available at FIGSHARE: https://doi.org/10.6084/m9.figshare.26953714.v1.

### Primary outcome measure – SDMT

An increase in total scores and correct responses in SDMT, indicates improved information processing speed. Both groups’ individual results (S1Appendix 1; Table 1) and ANOVA analysis (S2 Appendix 2; Tables 1–4) showed significant improvements from baseline to intervention (*p* < 0.001). The experimental group improved from mean (M) = 55.1, standard deviation (SD) = 13.8 to Μ = 61.3 (SD = 14.1), while the active control group increased from M = 55 (SD = 13.4) to M = 57.2 (SD = 13.2). A significant interaction between study phases and group (*p* < 0.001) indicated greater improvement in the experimental group. Baseline scores between study groups did not differ significantly (Fig 3), confirming their comparability. Post-hoc Bonferroni analysis and *t*–test confirmed significantly greater improvements (*t*(18) = 8.6, *p* < 0.05) in the experimental group (M = 6.2 (SD = 1.4)) compared to the active control group (M = 2.2 (SD = 0.6)).

**Fig 3.** Symbol Digit Modalities Test Performance Across Study Phases and Groups. *, Significant improvement (*p* ≤ 0.05) at the specified assessment point between the two study groups. The x-axis represents the study phases, while the y-axis displays the mean test scores. Both groups began with comparable baseline values (*p* = 1), thereby reducing the likelihood that initial differences influenced the observed outcomes. Although both groups showed improvements over time, the experimental group (white circles) demonstrated a significantly greater increase in performance compared to the active control group (black circles) (*p* < 0.05). These findings suggest that the in-phase bilateral upper limb exercise protocol may enhance cognitive processing more effectively than the control intervention.

**Table 3.** Correlations Between Improvements in Purdue Pegboard subtests and Symbol Digit Modalities Test, for the Experimental Group.

|  | SDMT | Dominant Hand | Non-Dominant Hand | Bimanual | Assembly |
| --- | --- | --- | --- | --- | --- |
| SDMT |  |  |  |  |  |
| Dominant Hand | $r = 0.6$<br>$p = 0.07$ | | | | |
| Non-Dominant | $r = 0.6$<br>$p = 0.09$ | $r = 0.8$<br>$p = 0.006 *$ | | | |

| Hand |  |  |  |  |  |
| --- | --- | --- | --- | --- | --- |
| **Bimanual** | $r = 0.5$<br>$p = 0.1$ | $r = 0.8$<br>$p = 0.01 *$ | $r = 0.9$<br>$p = 0.001 *$ | | |
| **Assembly** | $r = 0.7$<br>$p = 0.02$ | $r = 0.8$<br>$p = 0.007 *$ | $r = 0.9$<br>$p = 0.001 *$ | $r = 0.8$<br>$p = 0.006 *$ | |

SDMT, Symbol Digit Modalities Test; $r$ , Pearson correlation coefficient; $p$ , $p$ – value; \*, Significant after Bonferroni correction at $p \leq 0.05$ . In the experimental group non-significant moderate correlation was observed between the SDMT and the unimanual dominant ( $r = 0.6$ , $p = 0.07$ ), the unimanual non-dominant ( $r = 0.6$ , $p = 0.09$ ) and the bimanual ( $r = 0.5$ , $p = 0.1$ ) subtests. However, significant moderate correlation was observed between the SDMT and the Assembly subtest ( $r = 0.7$ , $p = 0.02$ ). The highest correlation was observed between a) the unimanual dominant and bimanual subtests ( $r = 0.9$ , $p = 0.001$ ) and b) the unimanual dominant and Assembly subtests ( $r = 0.9$ , $p = 0.001$ ).

**Table 4.**
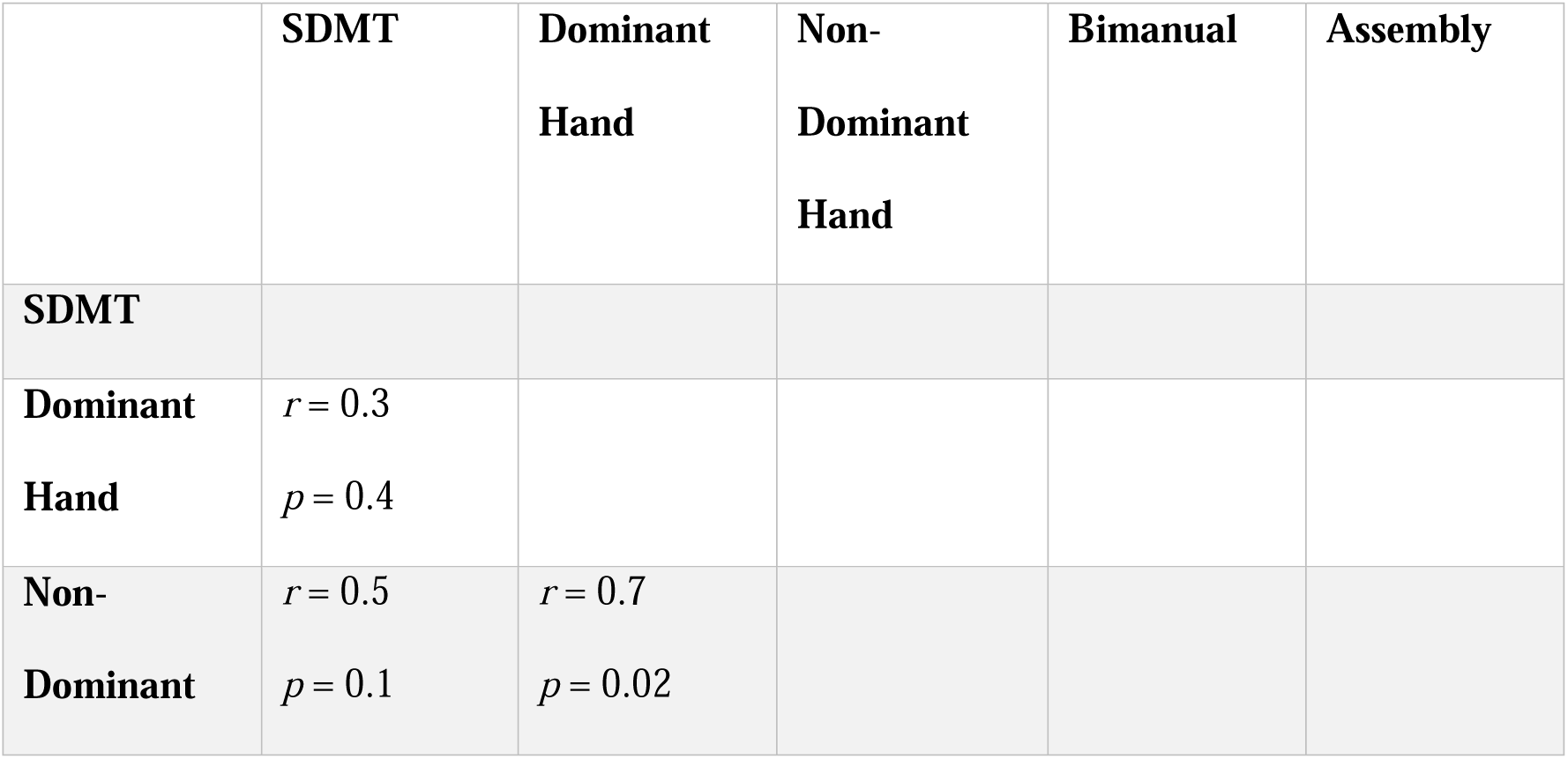
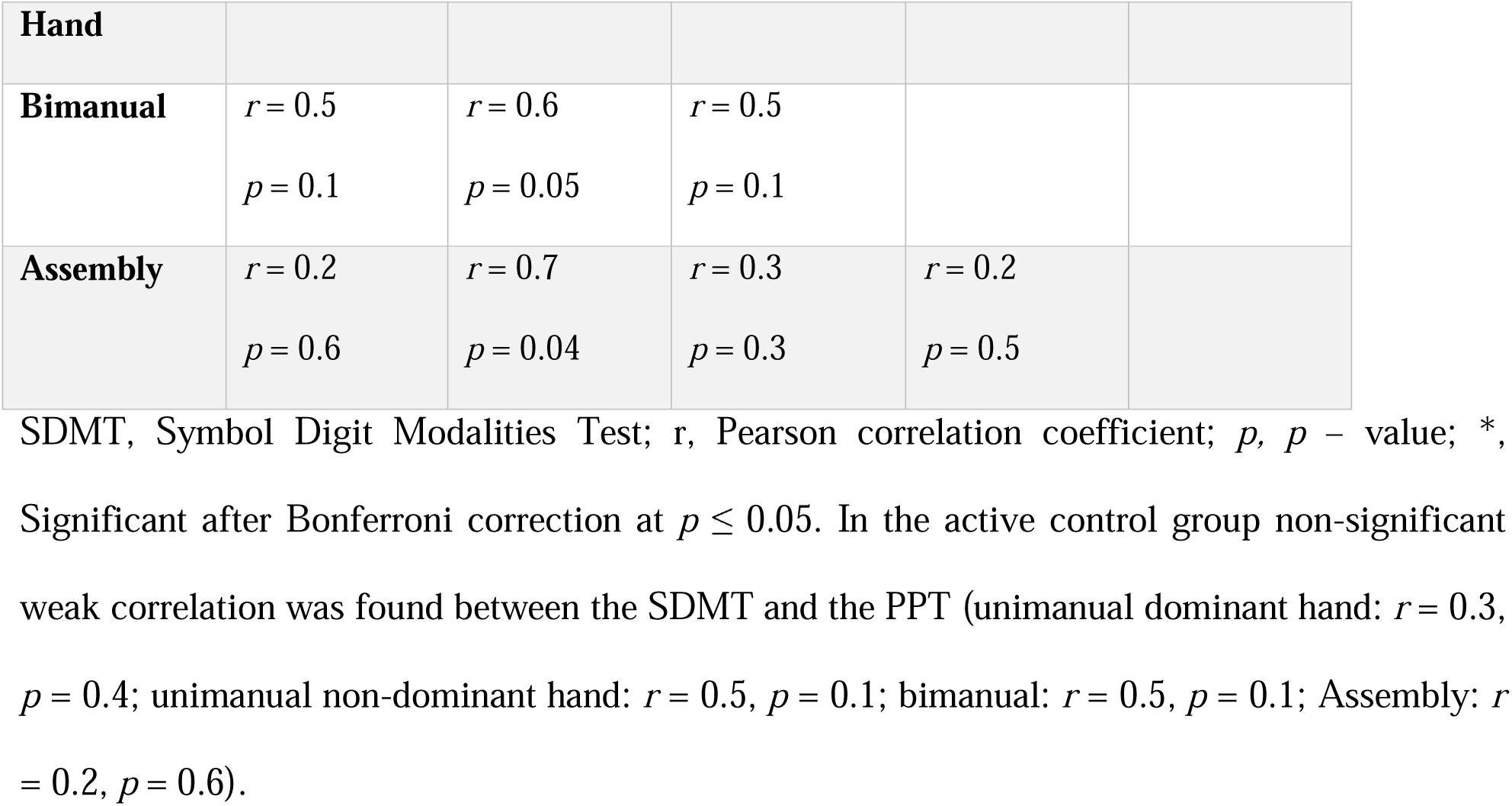
Correlations Between Improvements in Purdue Pegboard subtests and Symbol Digit Modalities Test, for the Active Control Group.

Focused on the primary outcome (i.e., SDMT), with an estimated effect size of partial η² = 0.807, a significance level of α = 0.05, and a sample size of 20 participants, a post hoc analysis indicated that the study had 100% statistical power to detect associations (1 – β = 1). The power analysis was conducted using G*Power (53).

These findings suggest that the in-phase bilateral upper limb exercise protocol may enhance cognitive processing more effectively than the conventional exercises.

### Secondary outcome measures

#### Medical Outcomes Study Questionnaire Short Form 36 Health Survey

Higher scores reflected better QoL. ANOVA analysis (S1 Appendix 1; Tables 5–8) and the individual results (S2 Appendix 2; Table 2) showed significant improvement in the experimental group from baseline (M = 88.8 (SD = 6.1)) to intervention (M = 95.6 (SD = 5.5); *p* = 0.029), with no change of the active control (baseline; M = 85.7 (SD = 8.3) to intervention; M = 85.6 (SD = 6.7). A significant group-by-phase interaction was observed (*p* = 0.009) and the post-hoc analysis confirmed within-group improvements in the experimental group only (Fig 4). An independent *t* – test further supported a greater improvement (*t*(18) = 7.03, *p* < 0.05) in the experimental group (M = 6.8 (SD = 2.3)) compared to the active control group (M = −0.06 (SD = 2.9)).

**Fig 4.** Performance of Medical Outcomes Study Questionnaire Short Form 36-Item Health Survey Across Study Phases and Groups. I, Intervention; *, Significant improvement (*p* ≤ 0.05) at the specified assessment point between the two study groups. The x-axis displays the assessment time points, including baseline (mean score) and three subsequent intervention phases (I1, I2, I3), while the y-axis represents the mean test scores for the experimental group (white circles) and the active control group (black circles). At baseline, there were no significant differences between groups (*p* = 1), indicating comparable starting points and minimizing the risk of baseline confounding. Over the course of the intervention, the experimental group exhibited a progressive and statistically significant improvement in test scores (I1: *p* < 0.001; I2: *p* < 0.001; I3: *p* = 0.013), whereas the active control group showed no improvement. These findings suggest that the in-phase bilateral upper limb exercise protocol led to a meaningful improvement in perceived quality of life in the experimental group.

#### Modified Fatigue Impact Scale

Lower scores indicated less fatigue. Fatigue significantly decreased in the experimental group only, with mean scores dropping from baseline M = 34 (SD = 14.8) to intervention M = 26.7 (SD = 15) (*p* < 0.001), while the active control group showed no change. ANOVA analysis (S2 Appendix 2; Tables 9 – 12) and the individual results (S1 Appendix 1; Table 3) confirmed a significant group-by-phases interaction (*p* < 0.001), with no baseline differences. Post-hoc tests (Bonferroni-corrected) showed significant improvements at all intervention time points, in the experimental group only (Fig 5). A *t–*test confirmed greater improvement in the experimental group (M = 10.6 (SD = 7.9)) versus active control group (M = −0.3 (SD = 1.6)), *t*(18) = 4.1, *p* < 0.05.

**Fig 5.** Performance of Modified Fatigue Impact Scale Across Study Phases and Groups. I, Intervention; *, Significant improvement (*p* ≤ 0.05) at the specified assessment point between the two study groups. The x-axis displays the baseline assessment (mean score) and three subsequent intervention time points (I1, I2, I3), while the y-axis represents the mean test scores for the experimental (white circles) and active control (black circles) groups. At baseline, the mean scores of the two groups did not significantly differ (*p* = 1), indicating comparable starting points and reducing the likelihood that initial differences influenced the observed outcomes. Over the course of the intervention, the experimental group exhibited a progressive and sustained improvement, reflected by a significant reduction in performance scores at each time point (I1: *p* = 0.004; I2: *p* < 0.001; I3: *p* < 0.001). In contrast, the active control group showed no significant change across the same period. These findings suggest that the in-phase bilateral upper limb exercise protocol led to a significant reduction in fatigue levels in the experimental group.

#### Purdue Pegboard Test

Higher scores reflected improved performance on the PPT, indicating enhanced manual dexterity. Both experimental and active control groups showed changes from baseline to intervention across all subtests (S1 Appendix 1; Tables 4 – 7). In the experimental group, mean scores increased in the Unimanual Dominant (baseline; M = 12.2 (SD = 2.3) to intervention; M = 14.2 (SD = 2.3)), Unimanual Non-Dominant (baseline; M = 10.3 (SD = 2) to intervention; M = 11.8 (SD = 2.2)), Bimanual (baseline; M = 9.4 (SD = 1.3) to intervention; M = 11.1 (SD = 1.7)) and Assembly (baseline; M = 17.6 (SD = 4.5) to intervention; M = 22.3 (SD = 5.4)) subtests, while the active control group exhibited minimal change (Fig 6).

**Fig 6.** Performance on the Purdue Pegboard Test Across Study Phases and Groups. Abbreviation: *, Significant improvement (*p* ≤ 0.05) at the specified assessment point between the two study groups. The mean assessment points per study phase are presented on the x-axis, whereas on the y-axis the mean scores are shown for each Purdue Pegboard subtest: Unimanual Dominant (A), Unimanual Non-Dominant (B), Bimanual (C) and Assembly (D), at baseline and intervention. The experimental group (white circles) demonstrated significant improvements across all subtests (all *ps* < 0.001), whereas this was not detected in the active control group (black circles), as indicated form the statistical analysis. Baseline characteristics of both groups, as indicated by their mean scores, showed no significant differences (all *ps* = 1), suggesting that both groups started from comparable baselines, minimizing the potential influence of initial differences on the outcomes. These findings suggest enhanced manual dexterity following the in-phase bilateral upper limb exercise protocol.

The ANOVA analysis (S2 Appendix 2; Tables 13 – 18) revealed significant effects of study phases and subtests (*p* < 0.001), with significant interactions 1) between study phases and group, 2) subtests and group (all *p*s = 0.035) and 3) a three-way interaction among phases, subtests and group (*p* < 0.001). Groups were equivalent at baseline (*p* = 1 for all subtests), confirming comparable starting points and reducing the likelihood that initial differences influenced outcomes. Bonferroni-corrected post-hoc tests showed significant improvements in the experimental group for all subtests except Unimanual Non-Dominant (*p* = 0.6), while no significant changes were observed in the active control group.

The greatest improvement in the experimental group occurred in the Assembly subtest (+4.6 pegs), significantly exceeding improvements in other subtests (*p* < 0.01) (Fig 7). No significant within-group differences were found among other subtests or in the controls (*p* = 1). Results indicate that the in-phase bilateral upper limb exercise protocol led to greater improvements in manual dexterity compared to the conventional exercise program.

**Fig 7.** Mean Improvement Scores Across Purdue Pegboard Subtests for the Experimental Group. The Purdue Pegboard subtests are shown on the x-axis, while the y-axis represents improvement measured by the number of pegs for each subtest. Bar plot showing average improvement in performance (baseline – intervention) across the four subtests of the Purdue Pegboard Test: Unilateral Dominant, Unilateral Non-Dominant, Bimanual and Assembly. The largest improvement was observed in the Assembly subtest (4.6 pegs) compared to the other subtests: Unimanual Dominant (2 pegs), Unimanual Non-Dominant (1.5 pegs), and Bimanual (1.7 pegs), indicating greater enhancement in complex, coordinated manual dexterity following the in-phase bilateral upper limb exercise protocol.

#### Timed 25-Foot Walk

Lower values indicated better gait performance. In the experimental group, gait performance improved significantly with test mean time decreasing from baseline M = 9 (SD = 2.3) to intervention M = 6.7 (SD = 2) (*p* < 0.001), unlike controls (baseline; M = 4.4 (SD = 2.7) to intervention; M = 9.5 (SD = 2.8)). ANOVA analysis (S2 Appendix 2; Tables 19 – 22) and the individual results (S1 Appendix 1; Table 9) from both study groups showed a significant group-by-phase interaction (*p* < 0.001). Post-hoc analysis and the *t–*tests confirmed greater improvement (*p* < 0.05) in the experimental group (M = 2.2 (SD = 0.7)) compared to the active control group (M = −0.05 (SD = 0.2)), with comparable baseline mean times (Fig 8).

**Fig 8.** Performance on the Timed 25-Foot Walk Test Across Study Phases and Groups. *, Significant improvement (*p* ≤ 0.05) at the specified assessment point between the two study groups. The x-axis represents the study phases, while the y-axis displays the mean test scores for the experimental (white circles) and active control (black circles) groups. Baseline comparisons between groups revealed no significant differences in mean scores (*p* = 1), indicating comparable starting points and reducing the likelihood that initial disparities influenced the outcomes. Over the course of the intervention, the experimental group exhibited a significant and sustained improvement (*p* < 0.001), reflected by a progressive decline in test scores. In contrast, the active control group showed no improvement. These findings suggest that the in-phase bilateral upper limb exercise protocol led to a significant enhancement in gait speed.

#### Six Spot Step Test

Lower values indicated better lower limb function. Test completion time significantly improved in the experimental group (baseline; M = 13.3 (SD = 8.3) to intervention; M = 11 (SD = 6.6), *p* = 0.022) based on non-parametric Wilcoxon signed-rank test), with no change in controls (baseline; M = 16.4 (SD = 8.7) to intervention; M = 16.5 (SD = 8.9)) (Fig 9). Individual data shown in S1 Appendix 1; Table 10.

**Fig 9.** Performance on the Six Spot Step Test Across Study Phases and Groups. *, Significant improvement (*p* ≤ 0.05) at the specified assessment point between the two study groups. The x-axis represents the study phases, while the y-axis displays the mean test scores for the experimental (white circles) and active control (black circles) groups. Baseline comparisons between groups revealed no significant differences in mean scores (*p* = 1), indicating comparable starting points and reducing the likelihood that initial disparities influenced the outcomes. Over the course of the intervention, the experimental group exhibited a significant and sustained improvement (*p* < 0.001), reflected by a progressive decline in test scores. In contrast, the active control group showed no improvement. These findings suggest that the in-phase bilateral upper limb exercise protocol led to a significant enhancement in gait speed.

### Correlation between SDMT and PPT

Pearson’s correlation coefficient (Pearson’s r) was calculated using the mean score from the intervention phase to examine the relationship between information processing speed and manual dexterity. The analysis included the SDMT and the sub-tests of the PPT. In the experimental group (Table 3) non-significant moderate correlation was observed between the SDMT and the unimanual dominant hand dexterity (r = 0.6, *p* = 0.07), the unimanual non-dominant hand dexterity (r = 0.6, *p* = 0.09) and the bimanual dexterity (r = 0.5, *p* = 0.1). Individual results for each test are presented in Fig 10 and in S1 Appendix 1 (Tables 1 and 4–7). However, significant moderate correlation was observed between the SDMT and the Assembly dexterity (r = 0.7, *p* = 0.02). On the other hand, in the active control group (Table 4) non-significant weak correlation was found between the SDMT and the Purdue Pegboard sub-tests (unimanual dominant hand: r = 0.3, *p* = 0.4; unimanual non-dominant hand: r = 0.5, *p* = 0.1; bimanual: r = 0.5, *p* = 0.1; Assembly: r = 0.2, *p* = 0.6). Nevertheless, the highest significant correlation was found in the experimental group, between two couplings of Purdue Pegboard sub-tests, the unimanual non-dominant hand - bimanual (r = 0.9, *p* = 0.001) and the unimanual non-dominant hand - Assembly (r = 0.9, *p* = 0.001). Following Bonferroni correction for five tests, statistically significant *p* – value is equal or less than 0.01 (*p* = 0.05 / number of tests = 5).

**Fig 10.** Individual and group results across Symbol Digit Modalities Test and Purdue Pegboard Test subtests. SDMT, Symbol Digit Modalities Test; PPT, Purdue Pegboard Test. Distribution of cognitive (i.e., SDMT) and manual dexterity test (i.e., PPT) scores across subtests for each group at baseline and intervention phases. Boxplots represent the median and interquartile range, while points show individual mean participant scores across the three baseline and intervention observations for (A) SDMT scores for the experimental group, (B) SDMT scores for the active control group, (C) PPT scores for the experimental group and (D) PPT scores for the active control group.

## Discussion

Exercise has increasingly been recognized as a comprehensive rehabilitation strategy capable of concurrently addressing both cognitive and motor domains (3,4), thereby offering broader therapeutic benefits than cognitive training alone (6). This pilot study sought to investigate the potential efficacy of in-phase bilateral upper limb exercises in pwPMS, yielding several novel findings. First, participation in a 12-week exercise protocol, based on in-phase bilateral upper limb movements, led to significant improvements in information processing speed, however, these cognitive gains did not correlate significantly with changes in manual dexterity. Second, the specific exercise protocol elicited marked improvements in balance, gait, fatigue and QoL when compared with the conventional exercise program.

### In-phase Bilateral Exercises – Information Processing Speed

Statistical analysis of the SDMT revealed significant improvements in both groups, indicating enhanced information processing speed in pwPMS (54,55), with significantly greater gains observed in the experimental group. As no participants engaged in structured exercise prior to the intervention, baseline cognitive function remained stable, suggesting that observed improvements were attributable to the intervention.

Information processing speed is a critical prognostic indicator of physical impairment (56) and represents the most prevalent cognitive deficit (3,4,57) in pwPMS (54,55). These findings align with prior research demonstrating the cognitive benefits of exercise in this population (58–62), including our previous study showing enhanced cognitive processing performance following in-phase bilateral upper-limb exercises in individuals with Relapsing-Remitting MS (63).

Emerging evidence supports a strong relationship between motor function and cognition in both healthy and MS populations (11,12,64–66). The anterior cingulate cortex, particularly its dorsal division, plays a central role in executive functions (i.e., attention, cognitive processing) (67,68) and is also activated during motor tasks (13,69,70). This region is a key node in motor-cognitive integration due to its dense projections to the motor cortex and spinal cord (13,15,69). Asemi et al. (2015) demonstrated that the dorsal anterior cingulate cortex modulates activity in the supplementary motor area, reinforcing its role in motor control (13). Additionally, bimanual coordination has been linked to enhanced intrahemispheric and transcallosal connectivity between the supplementary motor area and primary motor cortex (13,17,18,71). Grefkes et al. (2008) reported that in-phase bilateral upper-limb movements improve interhemispheric communication, supporting the cognitive-motor integration hypothesis (18). Based on our findings and existing evidence, the in-phase bilateral upper-limb exercise protocol enhances information processing speed in pwPMS, supporting its integration into neurorehabilitation.

### In-phase Bilateral Exercises – Manual dexterity

Our findings showed greater improvement in manual dexterity in the experimental group. Despite higher baseline scores, post-hoc analysis confirmed no significant between-group differences, indicating comparable initial performance across subtests.

Following the in-phase bilateral exercise protocol, the experimental group exhibited significantly greater improvements across all subtests, with the most notable gains in the Assembly task, the most cognitively demanding measure of manual dexterity (72,73). In contrast, the controls showed no significant changes. While improvements in the Unimanual Non-Dominant subtest did not reach statistical significance, the observed trend aligns with prior research supporting the transfer effects of bilateral training (74,75) and its role in reducing lateral asymmetries (76).

These improvements may involve corpus callosum-mediated mechanisms that enhance bimanual coordination (77) and facilitate unilateral motor performance (78). Studies by Seitz et al. (2004) and Smith & Staines (2010) demonstrated that in-phase bimanual training increases cortical activation and improves motor performance in both clinical and healthy populations (78,79). Given that lesions in the corpus callosum are common in people with MS and can impair bimanual coordination (80,81), our findings, consistent with those of Seitz and Smith & Staines, suggest that in-phase bilateral upper-limb exercises may enhance bimanual dexterity in pwPMS.

Previous studies have shown that task-specific and intensive training is crucial for inducing neuroplastic changes in the non-dominant limb. The 12-week in-phase bilateral exercise protocol with three sessions per week may have been insufficient, as longer and more frequent interventions are often required to produce measurable changes (82–84). Nonetheless, the observed bilateral benefits indicate that this approach may be particularly valuable for individuals with motor impairments, who frequently underuse the non-dominant or affected upper limb.

### Information Processing Speed – Manual Dexterity Correlation

A Pearson’s correlation analysis was performed between SDMT and PPT to define if the improvement in information processing speed influences manual dexterity. Therefore, a moderate correlation emerged, with statistical significance observed only between SDMT and the Assembly subtest (Table 3 and 4). This linear relationship was present in the experimental group but absent in the active control control. These results are consistent with prior findings in pwPMS (12,65,71), individuals with mild cognitive impairment and healthy older adults (71,85). Notably, the Assembly subtest, the most cognitively demanding task in the PPT, showed the greatest improvement among experimental participants. The Assembly task requires advanced cognitive control (72,73), including sequential planning and coordinated use of both hands within time constraints. This supports that in-phase bilateral upper limb exercises can simultaneously enhance cognitive processing and manual dexterity. Such behavioral associations were not observed in the active control group, which engaged in conventional exercises.

### In-phase Bilateral Exercises – Secondary Outcome Measures

Participants in the experimental group demonstrated significant improvements across all secondary outcomes compared with controls. Although the exercise protocol in the experimental group primarily targeted upper-limb function, its circuit-based design likely contributed to the observed enhancements in gait and balance (86,87), as it incorporated both gait and balance practice through transitions between exercises. Furthermore, significant reductions in fatigue and improvements in QoL were observed, as measured by the Modified Fatigue Impact Scale and the Medical Outcomes Study Short Form-36 Health Survey. These findings align with previous research linking exercise to improved cognition, manual dexterity and QoL in pwPMS (88–90), despite the commonly reported increase in fatigue perception that often accompanies neurological improvement in this population (91–94).

### Limitations

The present study highlights several methodological considerations that should be addressed in future research. First, the absence of a follow-up assessment limits the ability to evaluate the durability and reliability of the observed effects, as well as to capture potential long-term benefits for participants.

Second, the relatively small sample size, although appropriate for a pilot study aimed at generating preliminary data to inform future trials, may constrain the generalizability of the findings. To mitigate this limitation, twenty participants were recruited and randomly allocated (1:1) to either the experimental or active control group, allowing for robust interpretation of both within- and between-group effects. Moreover, based on the observed mean difference between groups, the sample achieved 100% statistical power to detect effects on the primary outcome measure (i.e., SDMT; information processing speed), thereby providing a strong empirical foundation for subsequent large-scale investigations.

Third, the unequal frequency of exercise sessions between groups may have affected the interpretation of the outcomes and underscores the necessity of adopting standardized training protocols in future investigations to enhance comparability. However, it should be noted that, as a pilot randomized controlled trial, the present study was primarily designed to evaluate the feasibility and preliminary efficacy of the in-phase bilateral intervention, rather than to delineate the specific components responsible for the observed effects. Consequently, the current study design does not permit differentiation of the effects attributable to exercise frequency and the findings should therefore be interpreted within an exploratory framework. Nevertheless, this study contributes to addressing a critical gap highlighted by De Luca et al. (2020), concerning the optimization of intervention dosage in people with MS (4).

Similarly, we acknowledge that, although the experimental group participated in a group-based exercise program, exercise rehabilitation is increasingly shifting toward individually prescribed protocols. Within this conceptual framework, group-based exercise in the present study was not implemented as an experimental variable, but rather as a strategy to enhance motivation, adherence and social engagement, factors known to positively influence cognitive outcomes. The differential effects of group-based versus individually delivered interventions, however, were beyond the scope of the current investigation.

Finally, the absence of neuroimaging or other qualitative outcome measures, such as functional magnetic resonance imaging, limits the ability to infer the neural mechanisms underlying the observed cognitive and motor improvements.

### Future Directions

To build upon the present findings, future research should replicate the exercise protocol in a larger cohort of pwPMS. Subsequent studies could also expand the methodological framework to incorporate quantitative neuroimaging assessments, enabling a more detailed investigation of the neural mechanisms underlying observed cognitive and motor changes. Moreover, the effects of in-phase bilateral exercises on additional cognitive domains, such as working memory and attention, should be systematically examined to delineate the broader cognitive benefits of this intervention. Finally, as group sessions were selected in the current pilot study to enhance social engagement and motivation, although the comparative effects of group versus individually delivered formats were beyond the study’s scope, future research should directly evaluate these delivery modes to clarify their distinct contributions to neurorehabilitation outcomes.

### Conclusion

Over recent decades, exercise has become a cornerstone in managing cognitive and motor impairments in pwPMS. Consistent with De Luca et al. (2020), who emphasized the need for holistic interventions targeting both domains (4), this pilot study examined the effects of in-phase bilateral upper-limb exercises on information processing speed in pwPMS. The group-based, circuit-format protocol was designed to concurrently engage cognitive and motor functions.

Preliminary findings suggest that in-phase bilateral training may enhance both cognitive processing and bimanual coordination, functions reliant on higher-order cognitive control (24,25,95). Given that in-phase bilateral exercises require less attentional load and motor control than unilateral exercises (25,26,96), the observed improvements in information processing speed and manual dexterity support their feasibility and potential as a targeted rehabilitation strategy in pwPMS. Although the current study was designed as a pilot randomized controlled trial with certain considerations, the findings provide preliminary evidence supporting the use of holistic, circuit-based, in-phase bilateral exercise program, combining sports activities and functional exercises, to enhance both motor and cognitive functions in pwPMS. These results warrant further investigation in larger, mechanistic trials to confirm efficacy and elucidate underlying neurophysiological mechanisms.

## Data Availability

Data are available at FIGSHARE: https://doi.org/10.6084/m9.figshare.26953714.v1.

## Acknowledgments

The authors gratefully acknowledge all individuals with multiple sclerosis who participated in this study. We also thank the Cyprus Sports Organization for providing access to the sports hall and a fitness instructor to support the intervention protocol, and in particular Dr. Elena Papacosta, board member, for her invaluable support. Generative AI (ChatGPT, OpenAI, version 2025) was used solely for language editing and grammatical refinement. The authors reviewed and approved all content and take full responsibility for the accuracy and integrity of the manuscript.

## Supporting Information

**S1 Appendix 1. Individual Results**

**S2 Appendix 2. ANOVA Analysis**

**S3 CONSORT 2025 checklist**

